# Interventions to help patients withdraw from depression drugs: systematic review

**DOI:** 10.1101/2023.03.13.23287182

**Authors:** Peter C. Gøtzsche, Maryanne Demasi

## Abstract

**BACKGROUND:** Depression drugs can be difficult to come off due to withdrawal symptoms. Gradual tapering with tapering support is needed to help patients withdraw safely. We reviewed the withdrawal success rates, using any intervention, and the effects on relapse/recurrence rates, symptom severity, quality of life, and withdrawal symptoms.

**METHODS:** Systematic review based on PubMed and Embase searches (last search 4 October 2022) of randomised trials with one or more treatment arms aimed at helping patients withdraw from a depression drug, regardless of indication for treatment. We calculated the mean and median success rates and the risk difference of depressive relapse when discontinuing or continuing depression drugs.

**RESULTS:** We included 13 studies (2085 participants). Three compared two withdrawal interventions and ten compared drug discontinuation vs. continuation. The success rates varied hugely between the trials (9% to 80%), with a weighted mean of 47% (95% confidence interval 38% to 57%) and a median of 50% (interquartile range 29% to 65%). A meta-regression showed that the length of taper was highly predictive for the risk of relapse (P = 0.00001). All the studies we reviewed confounded withdrawal symptoms with relapse; did not use hyperbolic tapering; withdrew the depression drug too fast in a linear fashion; and stopped it entirely when receptor occupancy was still high.

**CONCLUSIONS:** The true proportion of patients on depression drugs who can stop safely without relapse is likely considerably higher than the 50% we found.

## Introduction

Withdrawal symptoms can occur with all psychoactive drugs after prolonged exposure.^1,2^ For depression drugs, about half of the patients experience withdrawal symptoms when trying to stop or reduce the dose,^3^ and half of these rate the symptoms as severe.^3,4^ The symptoms are very diverse and may include flu-like symptoms, dizziness, shaking, fatigue, anxiety, bouts of crying, emotional lability, lowering of mood, electric shock sensations, akathisia, and even suicide, violence and homicide in rare cases.^5–7^ The withdrawal symptoms may persist for months or even years,^3^ and may mimic disease symptoms, which often results in doctors advising patients to resume drug treatment under the false assumption that the drug is necessary to prevent relapse. It is therefore crucial to successful and safe drug withdrawal that withdrawal symptoms are minimised by gradual tapering and are distinguished from relapse.

The incidence and severity of withdrawal symptoms depend on how the dose is tapered,^8–11^ and symptoms can occur even following very small dose reductions.^11,12^ After long-term exposure to a drug, the physiological adaptations take time to readapt back to normal. This leaves the affected receptors in a temporary disequilibrium following dose reduction, from which withdrawal symptoms arise.^1,11^

A drug should be tapered in a way that corresponds to a gradual and slow unblocking of its primary target receptor, which for most depression drugs is the serotonin transporter (SERT). The relationship between dose and SERT occupancy is not linear, but hyperbolic, and occupancy increases rapidly at lower doses and plateaus at moderate and higher doses.^13–17^

A linear tapering regimen corresponds to increasingly larger reductions in receptor occupancy as the tapering progresses and carries a high risk of withdrawal symptoms. Hyperbolic tapering is needed,^11,18^ involving numerous dose reductions below the lowest standard manufactured dose.

Coming off depression drugs can be difficult also for psychological reasons. As barriers and challenges for stopping depression drugs, patients point to anxiety, uncertainty, worry of relapse, insufficient emotion regulation skills, perceived etiology of depression being biochemical, and need for social support.^19–24^

The existence of peer-to-peer psychiatric drug withdrawal communities and survivor groups^25,26^ indicates an unmet need for helping patients withdraw from depression drugs.

Evidence-based guidance is needed to help patients who want to come off their drug. This includes optimal tapering and effective psychosocial support or therapy when needed. Unfortunately, official guidelines are often outright dangerous as they recommend fast tapering in a linear fashion.^3,7,27^

We reviewed trials of interventions to help patients withdraw from depression drugs. Our primary objective was to determine the success rate of patients attempting withdrawal from a depression drug, using any intervention. Our secondary objective was to determine the effects of interventions on relapse/recurrence rates, symptom severity, and quality of life. We also evaluated the applied tapering procedures and any education and information on withdrawal symptoms provided to the participants.

## Methods

We conducted a systematic review of the randomised trials^28^ with at least one treatment arm that aimed to help patients withdraw from a depression drug.

Eligible studies were those whose purpose was to investigate if stopping a depression drug was possible, in any language, and which reported on complete cessation of drug use. Crossover trials were excluded. We included two types of trials: those comparing two withdrawal interventions and those comparing one withdrawal intervention with continued maintenance drug treatment. For the latter, we were interested in “real-life” patients who wanted to withdraw. We therefore excluded studies where the drug was administered, and later discontinued, as part of the trial design to be compared with maintenance medication. We included relapse prevention trials where the experimental intervention involved discontinuing the patients’ current drug provided that complete cessation of drug use was measured.

Eligible participants were patients on any depression drug for any indication. Eligible interventions were any intervention aimed at helping patients withdraw.

Our primary outcome measure was the proportion of patients who succeeded to discontinue their depression drug. Drop-outs were categorized as unsuccessful discontinuation. Our secondary outcome measures were relapse/recurrence rates, symptom severity, quality of life, functioning, and withdrawal symptoms measured on any validated rating scale or interview.

We searched PubMed and Embase on January 11, 2021, and updated our searches on October 4, 2022. Search terms for PubMed: Antidepressant [MESH] and (discontinu* or withdraw* or taper* or stop* or cease* or cessat*). Search terms for Embase: exp antidepressant agent/ and (discontinu* or withdraw* or taper* or stop* or cease* or cessat* or deprescri*).af, limited to human, randomized controlled trial, and article or article in press. We scanned the references of retrieved articles and related review articles and contacted authors to obtain missing data.

Titles and abstracts were screened for eligibility by one researcher. Full texts of potentially eligible papers were read by two researchers. Disagreements were resolved by discussion. Reasons for exclusion after full-text reading were noted.

We used Zotero to manage the searches and Excel and Word for the extracted data. One researcher extracted data, and another researcher checked this, which led to a few minor changes. We used a standardised and piloted data extraction form and extracted data from the latest follow-up.

We calculated the weighted mean rate (with 95% confidence interval), and the median (with interquartile range) of successful withdrawal, counting dropouts as failures. For studies comparing two withdrawal interventions, we calculated the risk difference of successful withdrawal (with 95% confidence interval). We also calculated the risk difference of depressive relapse after discontinuing vs. continuing depression drugs. We used Comprehensive Meta Analysis (random effects model).

## Results

Our literature searches identified 4023 records from PubMed and 3431 from Embase initially and 164 and 325 new records, respectively, in our updated searches, a total of 7943 records. Many of these were duplicates or obviously irrelevant. After screening titles and abstracts, 40 articles remained for full text reading. We found an eligible study by scanning the references,^29^ and initially included 12 studies (13 publications)^29–41^ after having excluded ten single arm trials,^42–51^ four articles of interventions not aimed at withdrawal,^52–55^ three articles that were not trial reports,^56–58^ one article reporting the results at an earlier follow up,^59^ two retrospective observational studies,^60,61^ one study administering the depression drug as part of the trial,^62^ a crossover trial,^63^ and three studies that did not report withdrawal success rates.^64–66^ We obtained unpublished data from the author of one study.^32^ Our updated searches identified one additional eligible study listed with three publications in both databases.^67-69^

Three of the 13 included studies compared two withdrawal interventions^30–32^ and ten compared withdrawal with continued drug treatment (tables 1a and 1b).^29,33–40,67^ One study was prematurely stopped following an interim analysis conducted by an independent data monitoring committee due to excessive deterioration upon withdrawal, which was interpreted as relapse.^32^

**Table 1a.** Study characteristics of trials comparing two drug withdrawal interventions.

| Study | Withdrawal arm 1 (N) | Withdrawal arm 2 (N) | Diagnosis | Drug | Time on drug | Taper length | Dose reduction regimen |
| --- | --- | --- | --- | --- | --- | --- | --- |
| Fava 2004<sup>30</sup> | CBT + lifestyle change (23)<sub>a</sub> | Standard clinical care (22) | MDD in remission | TCA, SSRI | 3-5 mo | NA | Fixed, linear: 25 mg/2 wk (amitriptyline or similar) |
| Fava 1998<sup>31</sup> | CBT (21)<sub>a</sub> | Standard clinical care (22) | MDD in remission | Ami, des, imi, mia | 3-5 mo | NA | Fixed, linear: 25 mg/2 wk (amitriptyline or similar) |
| Scholten 2018<sup>32</sup> | CBT (42) | Standard clinical care (45) | Anxiety disorder in remission | NA | NA | 4 mo | Fixed, linear, every 2 wk |

**Table 1b.**
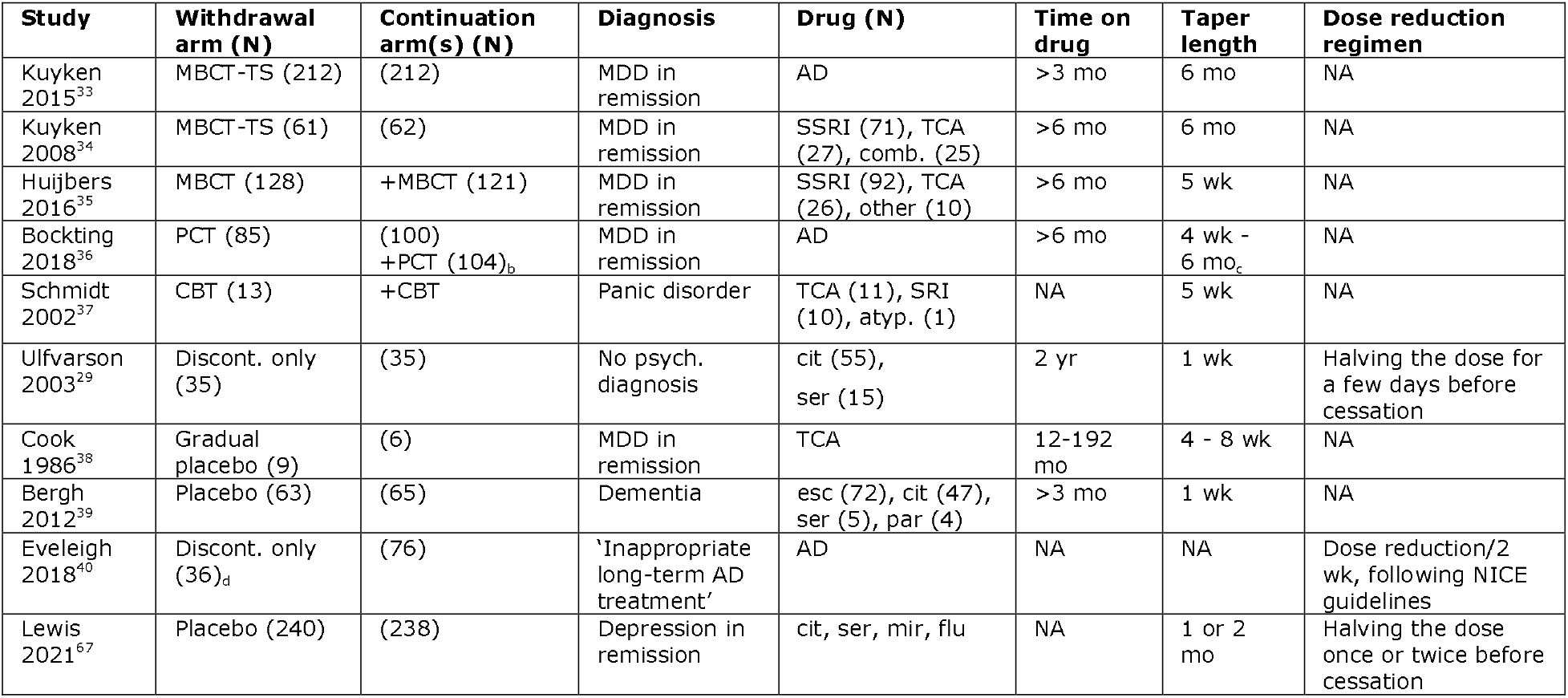
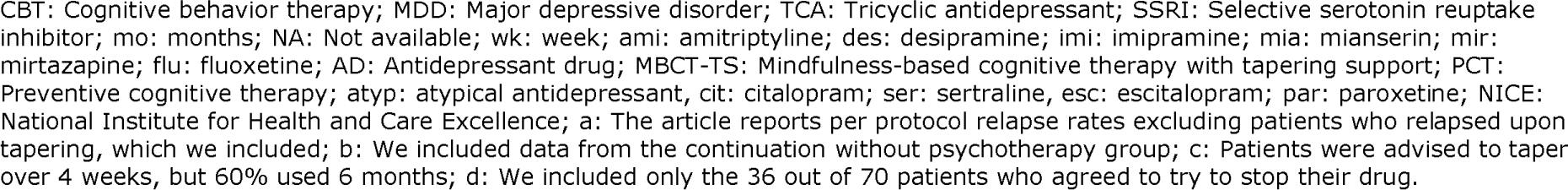
Study characteristics of trials comparing drug withdrawal with continuation.

| Study | Withdrawal arm (N) | Continuation arm(s) (N) | Diagnosis | Drug (N) | Time on drug | Taper length | Dose reduction regimen |
| --- | --- | --- | --- | --- | --- | --- | --- |
| Kuyken 2015<sup>33</sup> | MBCT-TS (212) | (212) | MDD in remission | AD | >3 mo | 6 mo | NA |
| Kuyken 2008<sup>34</sup> | MBCT-TS (61) | (62) | MDD in remission | SSRI (71), TCA (27), comb. (25) | >6 mo | 6 mo | NA |
| Huijbers 2016<sup>35</sup> | MBCT (128) | +MBCT (121) | MDD in remission | SSRI (92), TCA (26), other (10) | >6 mo | 5 wk | NA |
| Bockting 2018<sup>36</sup> | PCT (85) | (100) +PCT (104)<sub>b</sub> | MDD in remission | AD | >6 mo | 4 wk - 6 mo<sub>c</sub> | NA |
| Schmidt 2002<sup>37</sup> | CBT (13) | +CBT | Panic disorder | TCA (11), SRI (10), atyp. (1) | NA | 5 wk | NA |
| Ulfvarson 2003<sup>39</sup> | Discont. only (35) | (35) | No psych. diagnosis | cit (55), ser (15) | 2 yr | 1 wk | Halving the dose for a few days before cessation |
| Cook 1986<sup>38</sup> | Gradual placebo (9) | (6) | MDD in remission | TCA | 12-192 mo | 4 - 8 wk | NA |
| Bergh 2012<sup>39</sup> | Placebo (63) | (65) | Dementia | esc (72), cit (47), ser (5), par (4) | >3 mo | 1 wk | NA |
| Eveleigh 2018<sup>40</sup> | Discont. only (36)<sub>d</sub> | (76) | 'Inappropriate long-term AD treatment' | AD | NA | NA | Dose reduction/2 wk, following NICE guidelines |
| Lewis 2021<sup>67</sup> | Placebo (240) | (238) | Depression in remission | cit, ser, mir, flu | NA | 1 or 2 mo | Halving the dose once or twice before cessation |

CBT: Cognitive behavior therapy; MDD: Major depressive disorder; TCA: Tricyclic antidepressant; SSRI: Selective serotonin reuptake inhibitor; mo: months; NA: Not available; wk: week; ami: amitriptyline; des: desipramine; imi: imipramine; mia: mianserin; mir: mirtazapine; flu: fluoxetine; AD: Antidepressant drug; MBCT-TS: Mindfulness-based cognitive therapy with tapering support; PCT: Preventive cognitive therapy; atyp: atypical antidepressant, cit: citalopram; ser: sertraline, esc: escitalopram; par: paroxetine; NICE: National Institute for Health and Care Excellence; a: The article reports per protocol relapse rates excluding patients who relapsed upon tapering, which we included; b: We included data from the continuation without psychotherapy group; c: Patients were advised to taper over 4 weeks, but 60% used 6 months; d: We included only the 36 out of 70 patients who agreed to try to stop their drug.

The studies included 2085 patients, 1057 of whom were randomised to withdraw (175 in the trials comparing withdrawal strategies and 882 of 1910 in the discontinuation vs. continuation trials). The diagnosis was major depressive disorder in remission in seven studies,^30,31,33–36,38^ depression (ICD-10 criteria),^67^ anxiety disorder in remission,^32^ panic disorder,^37^ dementia,^39^ “inappropriate long-term antidepressant treatment,”^40^ and no psychiatric diagnosis (nursing home patients)^29^ in one study each. Four studies gave drug names,^29,31,39,67^ four studies the drug class^30,34,37,38^ and five studies just noted it was a depression drug.^32,33,35,36,40^ Treatment duration with the current drug was specified as the mean or range in four studies;^29–31,38^ five studies provided a minimum duration (ranging from >3 to >6 months);^33–36,39^ one the number of patients on drug for at least three years;^67^ and three studies provided no information.^32,37,40^

Tapering was assisted by cognitive behaviour therapy (CBT) in five studies^30–32,36,37^ and mindfulness-based cognitive therapy (MBCT) in three studies^33–35^ (two had added a specific tapering support component to the treatment manual: MBCTTS).^33,34^ Five studies discontinued the drug with no further support,^29,38–40,67^ three via placebo replacement.^38,39,67^ Three studies provided psychotherapy in both the withdrawal arm and the continuation arm,^35–37^ one of which was a three-armed study including also a maintenance only group.^36^

All studies but one^67^ appeared to have stopped the drug at the lowest standard manufactured dose – corresponding to high receptor occupancy – as there was no mention of splitting tablets or capsules into smaller units or of hyperbolic tapering. In the largest study,^67^ patients who were taking citalopram, sertraline, or mirtazapine received the medications at half their regular dose in the first month; the same dose and placebo on alternate days in the second month; and from then on, only placebo. Patients who were taking fluoxetine received 20 mg and placebo on alternate days in the first month and then only placebo “since fluoxetine has a long half-life.”^67^

In nine studies,^29,32–39^ the drug was tapered over a certain period of time, ranging from one week to six months, but seven of them did not provide any information on how the dose was reduced.^33–39^ Five studies specified this, all of which used a linear tapering regimen.^29–32,40^ A detailed tapering scheme was provided in two studies.^32,40^ In addition, three studies mentioned having used an individualized tapering scheme, but provided no details.^33,34,37^

### Cessation of drug use

The cessation success rates in the 13 studies (including a total of 16 withdrawal arms) varied hugely, between 9%^30^ and 80%^29^ (table 2), and was measured at widely different durations of follow-up (ranging from four weeks to six years).

**Table 2.**
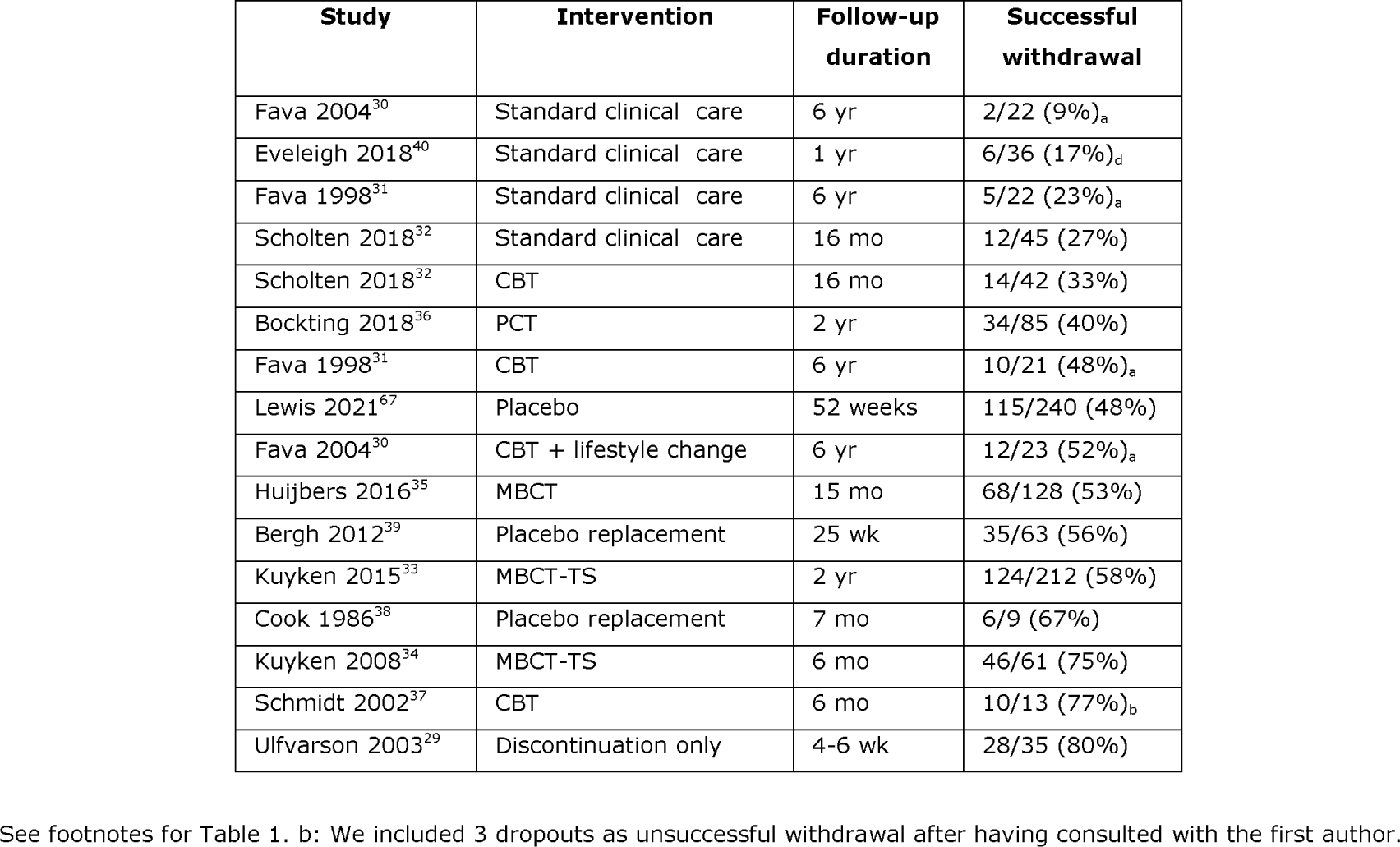
Success rates for patients attempting drug withdrawal.

The weighted mean success rate was 47% (95% confidence interval 38% to 57%, I^2^ = 90%, P < 0.0001). The median success rate was 50% (interquartile range 29% to 65%).

Offering CBT during tapering allowed significantly more patients to become drug free without relapse compared with tapering in standard clinical care (figure 1),^30–32^ risk difference 26% (95% CI: 10% to 43%, I^2^ = 41%, P = 0.002). Depression drug treatment duration before randomisation was unknown in one of the trials^32^ and rather short in the other two (3-5 months).^30,31^ The success rates were reported after six years^30,31^ and 16 months,^32^ respectively.

**Figure 1.**
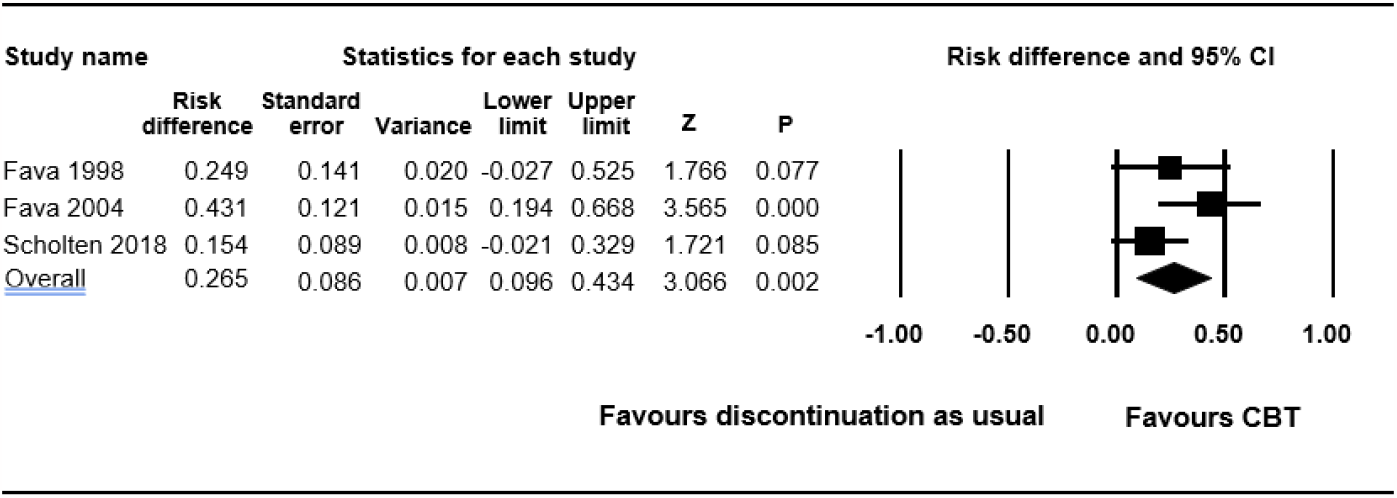
Rate of successful drug discontinuation using CBT vs. discontinuation as usual.

### Relapse of depression

Six trials compared relapse rates after discontinuing vs. continuing the drug in patients who were deemed in remission (figure 2). The risk of relapse was greater in the discontinuation group (risk difference 5.2%, 95% Cl: 0.4% to 10.1%, P = 0.035). However, there was substantial heterogeneity, (I^2^ = 79%), which may potentially be explained by the duration of the taper. In the three trials with results that favoured drug continuation, the drug was tapered over a markedly shorter period (4-8 weeks;^38^ 5 weeks;^35^ and 1-2 months,^67^ respectively) compared with the three trials that favoured discontinuation (6 months^33,34^ and 4 weeks to 6 months^36^). When we divided the trials into these two groups post hoc, the heterogeneity was zero for both analyses. There was no overlap between the two confidence intervals (figure 2), which suggests it is inappropriate to lump all six trials (P < 0.0001 for a comparison of the two estimates). The risk of relapse with short tapering was a risk difference of 17.4% (95% CI 10.4% to 24.4%) whereas it was -5.9% (−12.6% to 0.8%) with long tapering.

**Figure 2.**
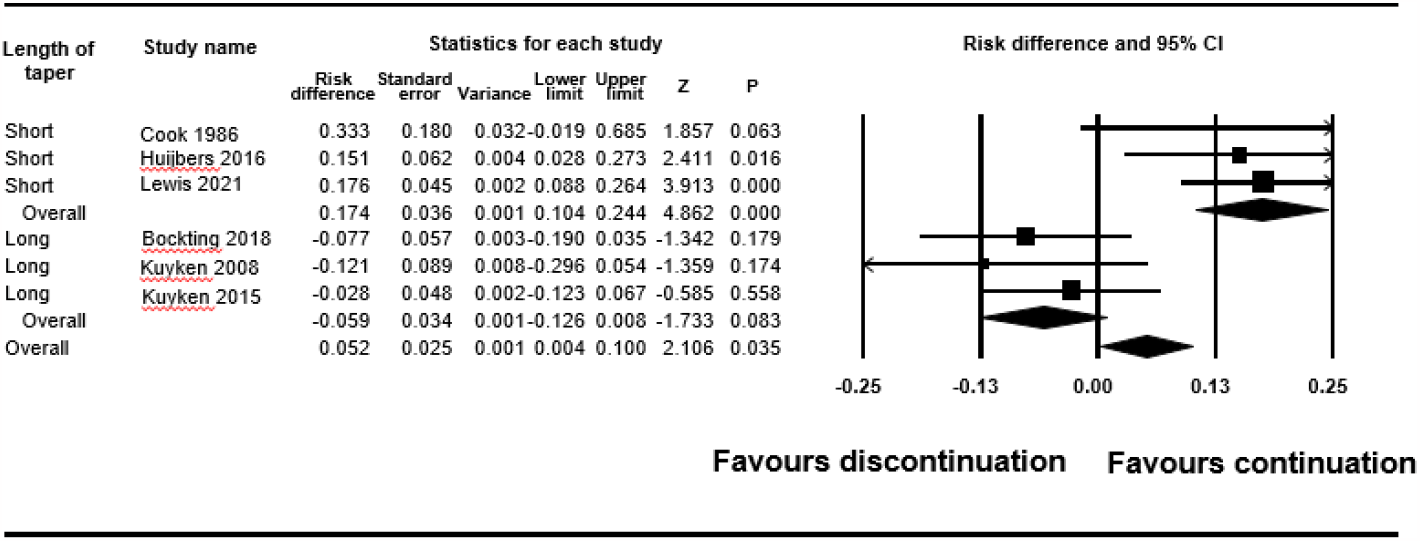
Rate of relapse of depression following drug discontinuation vs. continuation.

A peer reviewer was concerned about our analyses because her study, which declared a taper of 5 weeks in the publication,^35^ in practice had a taper with a median of 10 weeks. We therefore did a mixed effects meta-regression where we ranked the six studies 1 to 4 according to length of taper using 10 weeks for this study. It was highly significant (P = 0.00001), suggesting the length of taper is important for the risk of relapse (figure 3).

**Figure 3.**
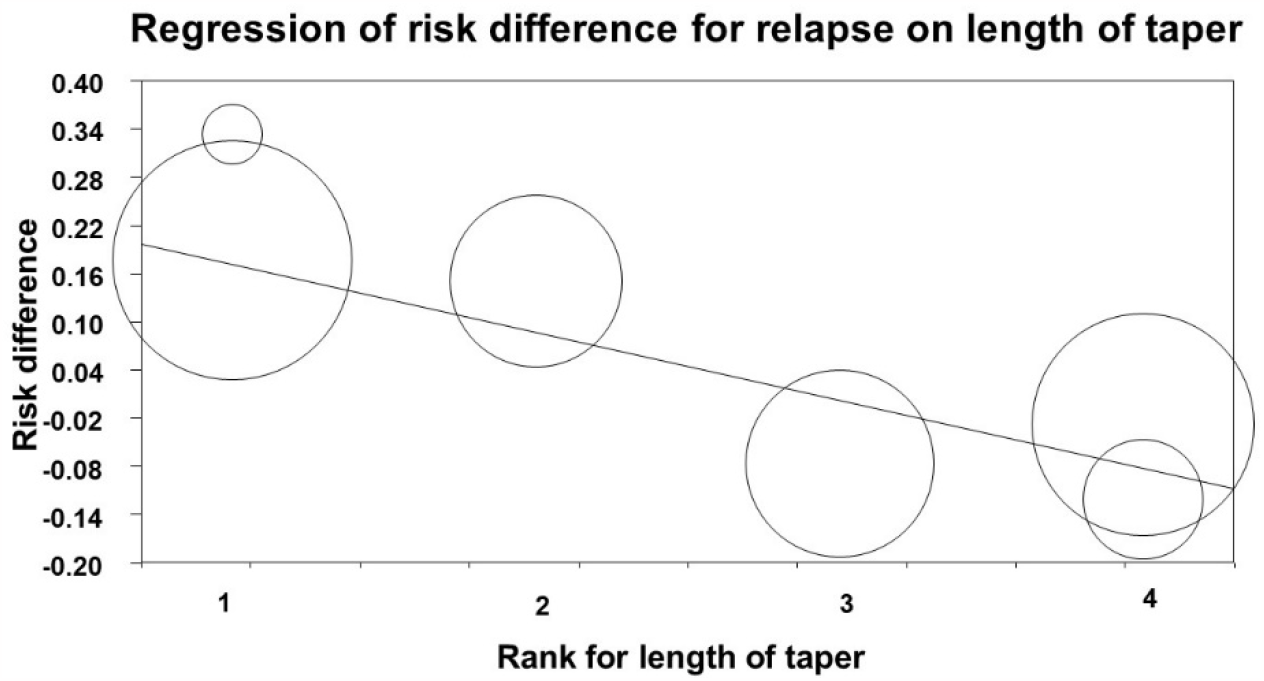
Meta-regression of the risk difference for relapse according to length of taper (P = 0.00001). The sizes of the circles reflect the weights of the studies. Studies: rank 1^38,67^; rank 2:^35^; rank 3:^36^; rank 4:^33,34^.

Out of the four trials using psychotherapy, the one that favoured drug continuation^35^ differed from the others by discontinuing the drug after therapy rather than during therapy; by administering psychotherapy (MBCT) to both groups; and by not having a specific tapering support component as the two other MBCT trials had. As this study was by far the largest of the two in the subgroup favouring drug continuation (accounting for 23% out of the total 31% weight in this subgroup), the observed heterogeneity between the subgroups may be explained by these differences.

### Quality of life, residual symptoms, and functioning

Other outcomes reported in the studies comparing discontinuation with continuation were quality of life, residual symptom severity, and functioning. Due to heterogeneity in participant population, diagnosis, rating scales, duration of follow-up, and degree and nature of psychological interventions, these outcomes could not be meta-analysed and are therefore presented narratively. Furthermore, mean differences and confidence intervals were rarely shown.

Quality of life appeared unchanged after stopping depression drugs compared with continued use in five trials (1349 patients)^29,33,35,39,67^ and improved in the psychological and physical domains, but not in the social domain, in one study with 123 patients.^34^

Symptom severity remained clinically unchanged in five studies (N=1243),^29,33,35,37,67^ improved on one of two scales in one study (N=123),^34^ and deteriorated in two studies (N=143; both of which used placebo substitution with no tapering support).^38,39^ The largest study reported selectively,^67^ as only results after 12 weeks in the 52-week study were described in the text. These results favoured continuation, in contrast to the 52-week results.

Functioning was measured in three studies, all of which remained unchanged after stopping the drug (N=220).^29,37,39^

### Suicide and suicidal ideation

None of the studies reported on suicidal ideation. One suicide occurred in the CBT + discontinuation group in the prematurely stopped trial.^32^ Two suicide attempts occurred; one during maintenance drug treatment and one in a patient who did not adhere to the tapering protocol but discontinued the drug abruptly.^36^

### Patients’ views

None of the studies reported on the patients’ views on the drug-free state or on the withdrawal process.

### Withdrawal symptoms

None of the studies reported on the incidence of withdrawal symptoms and no attempts at distinguishing between withdrawal symptoms and relapse were made. In three of the 13 studies, the authors considered the possibility of having misinterpreted withdrawal symptoms as relapse, but nevertheless concluded that deterioration *was* relapse.^32,35,39^ The remaining studies simply interpreted deterioration in the discontinuation group as relapse or recurrence.

## Discussion

Half of the patients succeeded in coming off their drug, and we found no convincing evidence of increased risk of relapse compared with continued maintenance drug treatment. The newest and largest study reported increased risk of relapse,^67^ but this was after a short and linear tapering regimen where the drug was discontinued at a dose corresponding to high receptor occupancy, introducing a high risk of confounding relapse with withdrawal symptoms.^17^

The studies found no difference in residual symptoms, functioning, and quality of life after stopping depression drugs. The effects of becoming drug free could not be isolated from the effects of the psychological interventions administered during withdrawal, most of which were aimed specifically at preventing relapse. However, most of the biases and uncertainties we identified favoured continued drug treatment.

These findings contrast with outcomes of traditional double-blinded discontinuation trials where the drug is stopped abruptly or rapidly, with no tapering support or psychotherapy, and with no distinction between withdrawal symptoms and relapse. Meta-analyses of such trials have been used to argue that continuing maintenance drug treatment reduces the risk of relapse by 50-70% compared with discontinuation.^70,71^ The studies we reviewed differ from such trials in several ways which could account for the contrasting findings: psychotherapy aimed specifically at relapse prevention was administered during tapering in the trials that found lower relapse rates in the discontinuation group; they had much longer tapering periods; and they included patients who wanted to stop.

Some of the heterogeneity in the 16 withdrawal arms in the studies we reviewed can be explained. The four lowest success rates (9-27%) occurred in groups using standard clinical care when tapering the drug and the success rate was closely related to the length of tapering.^30–32,40^

We found no indications that the patients were properly informed about type, incidence, duration, or possible severity of withdrawal symptoms.

Details on dose reductions and tapering regimens were very scarce, and most studies relied on tapering over a certain, fixed period. However, the duration of taper is largely irrelevant if the last dose reduction before cessation corresponds to a large drop in receptor occupancy.^11,17^ Receptor occupancy studies using PET and SPECT techniques consistently find that depression drugs are highly potent SERT antagonists even at the lowest available doses, where more than 50% of SERT are occupied.^15,17^

All six studies that specified the dose reductions followed a linear tapering regimen that was abruptly stopped when the dose was still high.^29–32,40,67^ The two detailed dose reduction schemes provided^32,40^ show that the receptor occupancy was a median 70%^17^ when the drug was stopped. In the largest trial, the fluoxetine dose before abrupt withdrawal was 10 mg,^67^ which corresponds to 73% receptor occupancy.^17^ It is highly likely that the outcomes reported in this trial were confounded by withdrawal symptoms.^72^ It is noteworthy, for example, that there was no difference in relapse rates the first month when the dose was still high in the discontinuation group, 50% of the starting dose.

As already noted, a gradual unblocking of SERT requires a hyperbolic reduction of dose,^11,17^ which involves performing multiple dose reductions below the lowest available doses. Hyperbolic tapering is laborious, as it involves splitting the smallest dose into smaller units, for example by using a nail file and a scale for tablets, dispersing the drug in water, counting beads for capsules,^7^ or using tapering strips.^73^ Clearly, if any study had done this, it would have been reported.

Due to the scarcity of randomised trials comparing different tapering rates and withdrawal interventions, we don’t know what the optimal tapering method is. But it is clear that the speed of tapering is highly individual.^7^

### Other systematic reviews of withdrawal

A systematic review from 2019 investigated interventions to facilitate discontinuation, but did not consider the tapering procedures as a relevant factor for successful withdrawal.^74^ Furthermore, study eligibility was restricted to adults with depression.

A 2021 review included only four trials,^75^ three of which we also included,^33,34,36^ whereas we excluded the trial that administered the drug as part of the trial.^62^

A 2021 Cochrane review on approaches for discontinuation versus continuation of long-term drug use did not include trials comparing different withdrawal strategies and it was restricted to adults with depression or anxiety.^76^ However, these drugs are being used for many conditions, and withdrawal symptoms are not dependent on why the drugs were prescribed. The Cochrane authors found that the risk of withdrawal symptoms may be similar in studies with rapid tapering schemes and in studies with abrupt discontinuation and that no firm conclusions can be made because relapse and other symptoms are confounded by withdrawal symptoms.

We initially planned to do a Cochrane review and submitted a protocol to the Cochrane Common Mental Disorders Group in 2017. The Cochrane group sent us on a 2-year mission that was impossible to accomplish, raising their demands along the way to absurd levels with many irrelevant requirements aimed at protecting the psychiatric guild.^77^ When the Cochrane group eventually refused to accept our protocol, it came with one of the worst peer reviews we have ever seen. This reviewer denied a long array of scientific facts we had presented and used several strawman arguments accusing us of things we had never claimed.^77^

The reviewer wanted us to “Start with a statement as to why antidepressants are considered by the scientific community to be beneficial … in treating a broad range of highly disabling and debilitating mental health problems.” We responded that our review was not an advertisement for the drugs and that it was not relevant to discuss their effect in a review about stopping using them. We were asked to explain the concept of ongoing prophylactic antidepressant treatment (maintenance therapy), “a well-accepted clinical strategy,” but this was outside the scope of our review and these trials are highly flawed because of withdrawal effects in the placebo group.^7,78^

We were required to accept and present the discarded hypothesis about depression being due to a chemical imbalance,^79^ and to mention that “some antidepressants may be more effective than others”, with reference to a network meta-analysis in *Lancet* by Cipriani and colleagues.^80^ This meta-analysis is deeply flawed.^81,82^ We were also criticised for our “stance on the relative harms and benefits of psychiatric drugs, which does not fully reflect the current international consensus and could cause alarm among review users who rely on Cochrane’s impartiality.”

One editor asked us to describe how antidepressants work and what the differences are between them, and a reviewer asked us to explain when it was appropriate and inappropriate to use antidepressants, but we were not writing a textbook in clinical pharmacology.

A reviewer wanted us to trivialise the harms by writing that, “some people get withdrawal symptoms that can negatively impact the quality of life of the patient.”

Meanwhile, we embarked on our review.

### Limitations in the trials

All the studies were seriously biased because withdrawal symptoms were confounded with relapse and other outcome measures.

The treatment duration before randomisation was rather short. In clinical practice, most patients are treated for many years,^83^ which makes it harder to withdraw.^84^ Conversely, all included patients were in remission, and since some real-life patients are unwell because of drug harms, they would likely benefit more from discontinuing the drug than patients who are well.

The withdrawal trials were not effectively blinded, which introduces a risk of bias.

Missing outcome data was an issue in several studies, with large proportions of patients dropping out, which was often unbalanced between the discontinuation group and the continuation group.

## Conclusion

All the studies we reviewed confounded withdrawal symptoms with relapse; did not use hyperbolic tapering; withdrew the depression drug too fast in a linear fashion; and stopped it entirely when receptor occupancy was still high. The true proportion of patients on depression drugs who can stop safely without relapse is likely considerably higher than the 50% we found.

## Data Availability

The data we have published are publicly available.

## Acknowledgments

We thank Willemijn Scholten for additional information on her trial.

## Availability of data

The data we have published are publicly available.

## Authors’ Note

Initially, the two researchers were Peter C Gøtzsche and the PhD student he had employed, psychologist Anders Sørensen. We submitted the review to a journal, which was very interested but asked for a revision. Sørensen promised to revise the manuscript but did nothing.

He did not respond to emails, never picked up the phone when he could see it was Gøtzsche who called and ignored telephone messages. After a year, Gøtzsche lost his patience and updated the literature search, added a new trial, responded to the peer review comments, and sent it all to Sørensen.

When Sørensen continued to ignore Gøtzsche, he asked the journal for advice. The editor suggested he drop Sørensen and add a new author, as there was a new trial to consider.

Gøtzsche submitted the revision with Maryanne Demasi. Then, the editor of the journal succeeded in making contact with Sørensen, who suggested his own changes, but sent them directly to the editor, without copying Gøtzsche or Demasi.

Gøtzsche added Sørensen’s name to the paper again, agreeing to most suggestions and resubmitted it to the journal. Then, again, Sørensen ignored all further emails from the journal, so we were instructed to publish without him, because the rules stipulate than an author must approve the final version.

The editor-in-chief asked Gøtzsche to get Sørensen’s signature confirming he was OK with not being an author. This was an impossible task, since Sørensen was now not responding to Gøtzsche or to the journal. We sought to ensure that Sørensen was in fact well, and we eventually established that he had been active with other projects.

The editor-in-chief got cold feet and asked the journal’s ethical team. After this, we were told they could not publish the paper.

The paper is highly important for psychiatric patients and for those who want to help them come off their drugs, which is part of Sørensen’s clinical practice.

It is unacceptable that a researcher is allowed to block publication of important research in the general interest. Since the standard is that researchers are free to publish independently if they cannot agree, we have decided to publish the review ourselves. A comment about our study and a link to it will be provided on the website of <u>Mad in America</u>, which is the obvious place to go to for those seeking reliable information about depression drugs.

